# Excessive monitoring for Hydroxychloroquine retinal toxicity in a trial to delay clinical type 1 diabetes created burden without benefit

**DOI:** 10.1101/2025.01.28.25321283

**Authors:** Sandra M Lord, Ingrid M Libman, Jane H Buckner, Kunal K Dansingani, Ken K Nischal, Polly J Bingley, Cate Speake, Lu You, Carla J Greenbaum

**Affiliations:** Benaroya Research Institute, Seattle, Washington, USA; Division of Endocrinology and Metabolism, Department of Medicine, University of Pittsburgh, Pittsburgh, Pennsylvania, USA; Department of Ophthalmology, University of Pittsburgh, Pittsburgh, Pennsylvania, USA; Division Chief Pediatric Ophthalmology and Strabismus; Medical Director Digital Health, UPMC Children’s Hospital of Pittsburgh; Assistant Medical Director UPMC International; Executive Vice Chair, UPMC Department of Ophthalmology; University of Pittsburgh, Pittsburgh, Pennsylvania, USA; Bristol Medical School, University of Bristol, UK; Health Informatics Institute, Morsani College of Medicine, University of South Florida, Tampa, FL, USA

## Abstract

**Background/Aims:** Type 1 Diabetes TrialNet (TrialNet) conducted a placebo randomized trial in children and adults with pre-clinical type 1 diabetes (T1D) to test whether hydroxychloroquine (HCQ) could delay or prevent progression to clinical T1D. Study regulators required more frequent monitoring for retinal toxicity than is recommended by clinical guidelines and we evaluated the impact of this monitoring.

**Methods:** We obtained the outcome of all eye exams from TrialNet and focused on exams that were recommended for follow up outside the protocol or for repeat testing within the trial.

**Results:** No HCQ related retinal toxicity was found, but increased frequency of monitoring led to additional testing due to incomplete exams or technical difficulties, and incidental findings prompted the need for follow up outside the study.

**Conclusion:** Our analysis suggests that increased frequency of retinal monitoring provided no benefit, created additional burdens, and that future trials testing HCQ in a pre-clinical T1D population could safely follow standard clinical guidelines for retinal surveillance.

## Background/Aims

HCQ is used to treat malaria, systemic lupus erythematosus (SLE), rheumatoid arthritis (RA), and juvenile idiopathic arthritis and there is increasing data to support its use in pre-clinical autoimmunity. The most important side effect of HCQ is retinal toxicity which is associated with cumulative exposure, calculated from dose and duration of use. The negligible incidence of retinal toxicity with less than five years of use informs current guidelines for retinal surveillance, which recommend a baseline eye exam to rule out existing pathology followed by annual exams after five years of use ^1^.

Current HCQ guidelines align with the low risk of retinal toxicity in the context of known benefit for clinical disease. However, there is no guidance about monitoring for retinal toxicity in pre-clinical disease, for which benefit is less clearly established. From 2018 - 2022, TrialNet tested HCQ in an asymptomatic population of children and adults with early stage (pre-clinical) T1D to determine whether T1D disease progression could be delayed or prevented ^2^. While the primary outcome of this study was negative, the frequency of retinal monitoring required by study regulators was more than standard clinical monitoring. Here, we evaluated the impact of frequent retinal monitoring in a population without clinical disease.

TrialNet is an NIH-sponsored international clinical trial network aiming to prevent, delay and reverse the progression of T1D. T1D is an autoimmune disease which proceeds from measurable autoantibodies in asymptomatic individuals to clinically apparent illness, characterized by hyperglycemia requiring life-long insulin therapy. Virtually all individuals with multiple autoantibodies will progress to clinical disease. The pre-clinical period is codified as early stages of T1D; Stage 1 T1D is defined as two or more pancreatic autoantibodies and normal glucose tolerance and Stage 2 T1D defined by progression to abnormal glucose tolerance ^3^.

HCQ has been shown to abrogate progression of palindromic rheumatism and to delay the onset of SLE in antibody positive individuals ^4-6^. Its immune effects include decreased T and B cell activation and signaling, reduced inflammatory cytokine production, and reduced autoantibody production ^7, 8^. HCQ may have positive effects on insulin sensitivity and beta cell function as supported by the reduced incidence of diabetes in individuals with SLE or RA who are treated with HCQ ^9, 10^. The combination of immune and metabolic effects suggests that HCQ is a potential therapy to delay clinical T1D.

## Methods

TrialNet enrolled 273 participants in the TrialNet Hydroxychloroquine Prevention study a 2:1 placebo randomized trial of HCQ in individuals aged 3-45 with pre-clinical (stage 1) T1D. Institutional or central review board approval was obtained at each site that participated in the Hydroxychloroquine in Stage 1 Type 1 Diabetes trial. Written informed consent was obtained from participants or their parent/legal guardian/next of kin to participate in the study. A majority of this study occurred during the COVID-19 pandemic. Twenty-two (15 in the HCQ arm, 7 in the placebo arm) individuals withdrew or were lost to follow-up within one month of enrollment. The study included 15 centers in the US and six international sites in Australia, Canada, Finland, Italy, UK, and Sweden. Study participants were randomly assigned to daily HCQ 5mg/kg/day to a maximum dose of 400mg/day or placebo. Participants were to undergo baseline (within 3 months of randomization) fundus photography, then annual visual field testing (VF) and spectral-domain optical coherence tomography (OCT) testing. Anticipating the challenges of testing young children, the protocol allowed for continuation of study drug for up to five years without complete OCT and VF exams, if testing was attempted annually. Photographs and scans were read and interpreted centrally by a single reader, or by a panel of ophthalmologists if the interpretation was uncertain. For this analysis, we obtained the outcome of all exams and focused on those recommended for follow-up outside the protocol or for repeat testing within the trial.

## Results

No study participants developed retinal toxicity over the median 23.3 months of observation ^2^. Forty-six individuals received HCQ for 3-4 years without retinal effects. Table 1 shows the number of individuals tested and test outcomes each visit. Baseline tests were performed up to three months after randomization on 243 individuals. Of these, 14 individuals (5.7%) had incidental findings, and an additional 36 (14.8%) individuals had incomplete or technically inadequate tests. There were 147 individuals who underwent scheduled tests at 12 months; 105 at 24 months; and 39 at 36-48 months. These follow-up tests revealed a further three incidental findings and 74 individuals had test results that were confounded by technical issues or were incomplete. Subjects with incidental findings were recommended for follow-up outside of the study. These findings included possible drusen; tortuous vessels raising suspicion of intracranial malformation; “saucerized” optic disc raising concern for glaucoma; indistinct optic disc margins raising concern for intracranial hypertension; choroidal nevus to be monitored for malignant transformation; vitreous cortex cells on OCT raising concern for uveitis. Study participants whose exams were confounded by technical issues or were incomplete were recommended for repeat testing and/or required additional attention from the study team. Two tests -- a 24-month OCT in an 18-year-old HCQ recipient and a 12-month VF in a nine-year-old placebo recipient--led to the recommendation to withhold the study drug pending panel review. In both cases, subsequent re-testing was normal.

**Table 1.** number of individuals tested and test outcomes each visit.

|  | Baseline – 3 months | 12 months | 24 months | 36 + months |
| --- | --- | --- | --- | --- |
| Subjects tested | 243 | 147 | 105 | 39 |
| Subjects with normal results | 193 (79.4%) | 110 (74.8%) | 70 (66.7%) | 32 |
| Subjects with incidental findings | 14 (5.7%) | 2 (1.3%) | 1 (0.9%) | 0 |
| Subjects with incomplete results or technical issues | 36 (14.8%) | 34 (23.1%) | 33 (31.4%) | 7 |
| Subjects with abnormal results-stop study drug, refer to panel | 0 | 1 (HCQ)<br>0.7% | 1 (placebo)<br>0.9% | 0 |

## Conclusions

There are guidelines to direct frequency of retinal exams in children and adults treated with HCQ for clinical conditions such as RA and SLE based on incidence of < 2% after 10 years of treatment ^11^. However, there is limited data to guide retinal surveillance in asymptomatic individuals with pre-clinical disease in a clinical trial setting, where benefit of HCQ is not established. The TrialNet HCQ for early stage T1D study found no evidence of HCQ induced retinopathy after a median of almost two years of exposure, and 46 individuals received HCQ for 3-4 years without retinal findings. The monitoring required in this trial was more frequent compared to standard clinical guidelines and was burdensome to sites and study participants. It led to repeated testing for incomplete exams or technical issues in up to 31% of subjects. Assessments in this healthy population also noted incidental findings, prompting referral outside the study, an additional financial and emotional hardship for subjects and their parents. While our analysis is limited in number, it suggests that individuals with pre-clinical T1D have a negligible risk of HCQ associated retinal toxicity and that future trials for which there is a prospect of direct benefit of HCQ therapy could safely follow standard clinical guidelines for retinal surveillance.

## Data Availability

All data produced in the present study are available upon reasonable request to the authors

## References

1. Rosenbaum JT, Costenbader KH, Desmarais J, et al. American College of Rheumatology, American Academy of Dermatology, Rheumatologic Dermatology Society, and American Academy of Ophthalmology 2020 Joint Statement on Hydroxychloroquine Use With Respect to Retinal Toxicity. Arthritis Rheumatol 2021 Jun;73(6):908–911.

2. Libman I, Bingley PJ, Becker D, et al. Hydroxychloroquine in Stage 1 Type 1 Diabetes. Diabetes Care. 2023 Nov 1;46(11):2035–2043.

3. Insel RA, Dunne JL, Atkinson MA, et al. Staging presymptomatic type 1 diabetes: a scientific statement of JDRF, the Endocrine Society, and the American Diabetes Association. Diabetes Care. 2015 Oct;38(10):1964–74.

4. Gonzalez-Lopez L, Gamez-Nava JI, Jhangri G, et al. Decreased progression to rheumatoid arthritis or other connective tissue diseases in patients with palindromic rheumatism treated with antimalarials. J Rheumatol. 2000 Jan;27(1):41–6.

5. Hanonen P, Möttönen T, Oka M. Treatment of palindromic rheumatism with chloroquine. Br Med J (Clin Res Ed). 1987 May 16;294(6582):1289.

6. James JA, Kim-Howard XR, Bruner BF, et al. Hydroxychloroquine sulfate treatment is associated with later onset of systemic lupus erythematosus. Lupus. 2007;16(6):401–9.

7. Katz SJ, Russell AS. Re-evaluation of antimalarials in treating rheumatic diseases: re-appreciation and insights into new mechanisms of action. Curr Opin Rheumatol. 2011 May;23(3):278–81.

8. Fox RI. Mechanism of action of hydroxychloroquine as an antirheumatic drug. Semin Arthritis Rheum. 1993 Oct;23(2 Suppl 1):82–91.

9. Solomon DH, Massarotti E, Garg R, et al. Association between disease-modifying antirheumatic drugs and diabetes risk in patients with rheumatoid arthritis and psoriasis. JAMA. 2011 Jun 22;305(24):2525–31.

10. Chen YM, Lin CH, Lan TH, et al. Hydroxychloroquine reduces risk of incident diabetes mellitus in lupus patients in a dose-dependent manner: a population-based cohort study. Rheumatology (Oxford). 2015 Jul;54(7):1244–9.

11. Melles RB, Marmor MF. The risk of toxic retinopathy in patients on long-term hydroxychloroquine therapy. JAMA Ophthalmol. 2014 Dec;132(12):1453–60. Erratum in: JAMA

